# Modelling trachoma post 2020: Opportunities for mitigating the impact of COVID-19 and accelerating progress towards elimination

**DOI:** 10.1101/2020.10.27.20219618

**Authors:** Anna Borlase, Seth Blumberg, E Kelly Callahan, Michael S Deiner, Scott D Nash, Travis C Porco, Anthony W Solomon, Thomas M Lietman, Joaquin M Prada, T. Dèirdre Hollingsworth

**Affiliations:** Big Data Institute, Li Ka Shing Centre for Health Information and Discovery, University of Oxford, Oxford, United Kingdom; Francis I Proctor Foundation, UCSF, USA; Trachoma Control Program, The Carter Center, Atlanta, Georgia, USA; Department of Control of Neglected Tropical Diseases, World Health Organisation, Geneva, Switzerland; Faculty of Health and Medical Sciences, University of Surrey, UK

**Keywords:** COVID-19, Elimination, Modelling, Trachoma

## Abstract

**Background:** The COVID-19 pandemic has disrupted planned annual antibiotic mass drug administration (MDA) activities which have formed the cornerstone of the largely successful global efforts to eliminate trachoma as a public health problem.

**Methods:** Using a mathematical model we investigate the impact of interruption to MDA in trachoma-endemic settings. We evaluate potential measures to mitigate this impact and consider alternative strategies for accelerating progress in those areas where the trachoma elimination targets may not be achievable otherwise.

**Results:** We demonstrate that for districts which were hyperendemic at baseline, or where the trachoma elimination thresholds have not already been achieved after 3 rounds of MDA, the interruption to planned MDA could lead to a delay greater than the duration of interruption. We also show that an additional round of MDA in the year following MDA resumption could effectively mitigate this delay. For districts where probability of elimination under annual MDA was already very low, we demonstrate that more intensive MDA schedules are needed to achieve agreed targets.

**Conclusion:** Through appropriate use of additional MDA, the impact of COVID-19 in terms of delay to reaching trachoma elimination targets can be effectively mitigated. Additionally, more frequent MDA may accelerate progress towards 2030 goals.

## Introduction

In response to the global COVID-19 pandemic, on 1 April 2020 the World Health Organization (WHO) released interim guidance that community-based surveys, active case-finding and mass drug administration (MDA) programmes for neglected tropical diseases (NTDs), including trachoma, be postponed.^1^

Annual district-level MDA of oral azithromycin, which targets the whole community, has formed the mainstay of global efforts to eliminate trachoma as a public health problem, with a single dose demonstrated to have good efficacy against ocular strains of *Chlamydia trachomatis*, the causative agent of trachoma.^2^ This strategy has proved to be widely successful in community-randomized studies and confirmed by the growing number of previously endemic countries which are now reaching the active trachoma threshold for elimination as a public health problem (EPHP), which is a prevalence of trachomatous inflammation—follicular (TF) in children aged 1-9 years of less than 5% (TF_1-9_<5%).^3–6^Given the pivotal role of annual MDA in the progress made in recent years, there are growing concerns regarding the potential impact of interruption to programmatic activities due to COVID-19. Upon cessation of MDA, empirical studies have indicated that in some areas infection returns exponentially, with a rate of resurgence anticipated to be faster in higher-transmission settings.^7,8^ In the context of programmatic activities being temporarily halted due to COVID-19, this is especially concerning for previously high-prevalence districts mid-way through trachoma elimination programmes. Furthermore, there are a finite number of districts where models suggest that under a strategy of annual MDA, elimination is not achievable,^9,10^ with the reproductive number under annual treatment (*R*_*T*_) being greater than 1 (defined as “MDA super-critical” by Blumberg and colleagues).^9^ For these districts there is concern that the interruption to programmes may cause a particularly marked surge in transmission with unknown consequences for morbidity. Regardless of the impact of COVID-19, these districts may need to implement alternative control strategies if EPHP is to be achieved.^9^

Building on previously developed mathematical models for trachoma transmission, we explore the impact of an interruption to community-level MDA in a range of endemic settings. Our individual-based stochastic model incorporates some key aspects of ocular *C. trachomatis* infection biology, including acquired immunity leading to decreased duration of infection with repeated infection and allows simulation of some of the variability in response to MDA observed in empirical studies.^2,7,11,12^ We also consider the effect of additional rounds of MDA in the year following the resumption of activities as a potential mitigation strategy. For those settings where reaching the EPHP target may not be achievable under current practice, we then explore the potential effectiveness of enhanced MDA protocols beyond 2020, as possible strategies for both mitigation and acceleration towards EPHP targets.

## Methods

### Model structure

The model for *C. trachomatis* transmission is based on a previously described framework,^13^ which accounts for TF persisting after clearance of ocular *C. trachomatis* infection. Individuals transitioning through four sequential states: Susceptible (S), infected but not yet diseased (I), infected and diseased (ID) or diseased but no longer infected (D), illustrated in Figure 1. Here disease refers to active trachoma, specifically TF. Within this framework, people who have cleared infection but remain diseased (D) are susceptible to infection but with force of infection (*λ*) reduced by a factor (*Г*). Further model description and model parameter definitions, values and sources are given in supplementary data (see Table S1).

**Figure 1.**
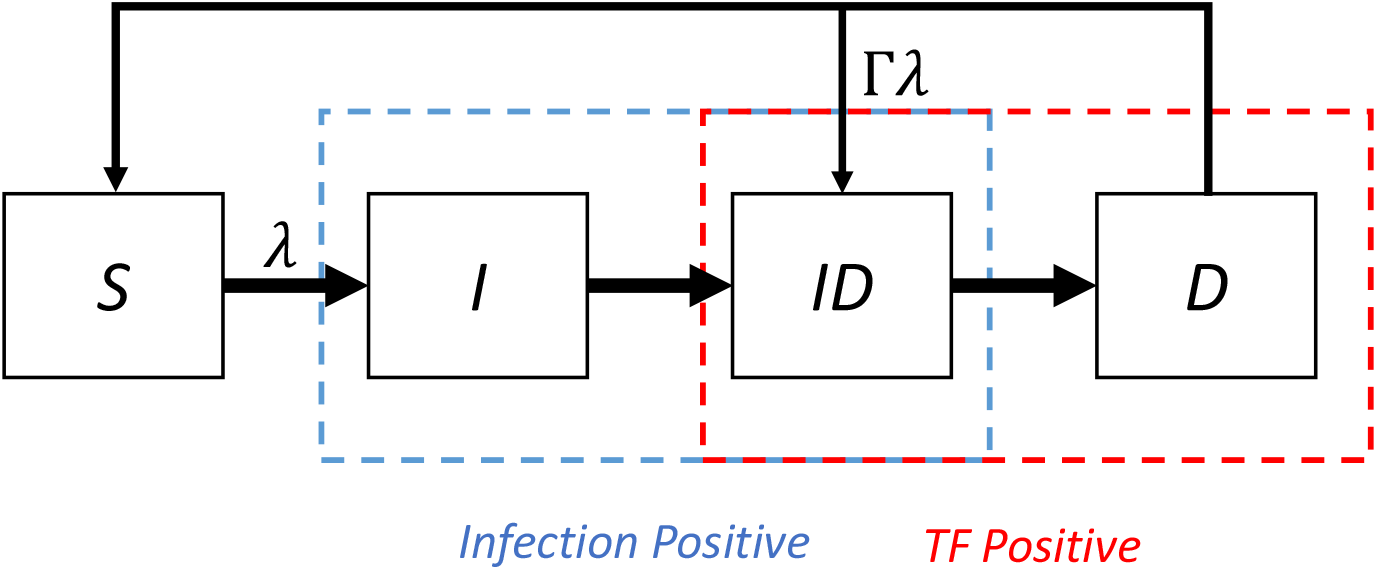
Schematic of model structure. Individuals can be susceptible to infection (S), infected but not yet diseased (I), infected and diseased (ID) or diseased but having cleared infection (D), where disease refers to trachomatous inflammation—follicular (TF). Individuals for whom infection has been cleared but disease persists (D) can be re-infected with force of infection (*λ*) reduced by *Г*.

The original ODE population-based model has previously been adapted to be stochastic, and has been further developed here to a fully stochastic individual-based model.^14^ Age, current infection/disease status and total number of infections for each individual are explicitly incorporated; the model runs in one-week timesteps.

Following the specifications of the original model and evidence from empirical studies, ^2,12,13^ duration of *ID* and *D* disease for each individual are assumed to decrease with each subsequent infection following a negative exponential; calculated durations for each infection are used as fixed transition periods in contrast to the exponential transitions utilised in previous models.^13,14^

## Treatment and systematic non-adherence

Community-wide MDA is assumed to be delivered to all ages with an 80% coverage level, in line with WHO minimum target coverage,^15^ and an efficacy (the probability that an individual who receives MDA clears infection) of 85% is assumed.^16^ To simulate the potentially lower efficacy of topical tetracycline eye ointment (which is routinely given to children aged less than 6 months), treatment is assumed to be 50% less effective in this age group.

In order to account for the possible role of systematic non-adherence to MDA, the “controlled correlation” method proposed by Dyson and colleagues is incorporated into the model (described in Supplementary Data).^17^ Briefly, *ρ* is the correlation parameter; if *ρ* =0, this is the equivalent to all rounds being randomly distributed (no systematic non-adherence) and if *ρ* =1 this corresponds to only those individuals who received the first round of treatment receiving future rounds (i.e. the same people are missed at every round; complete systematic non-adherence).

### Settings and Simulated Scenarios

#### Impact of 12 month interruption

Trachoma-endemic settings are generally categorised as hypo-, meso- and hyperendemic, corresponding to TF prevalence in ages 1-9 (TF_1-9_) of 5-9.9%, 10-29.9% and ≥30%. We modelled the different settings by varying the transmission parameter, with simulations fitted to prevalence category at most recent survey. Here we considered three alternatives: the most recent prevalence estimate could be either at baseline (i.e. before MDA has taken place), or at an impact survey after three or five rounds of annual MDA.

The average impact of a 12-month programme interruption (i.e. MDA is delivered 12 months later than intended, equivalent to a 2 year gap between MDA rounds) is then estimated in terms of average delay to reaching the threshold of TF_1-9_ <5%. The median delay is estimated as the difference between the median time to reaching the EPHP threshold after a 12 month interruption, and the median time to reaching the EPHP threshold if the interruption had not occurred (see Supplementary Data Table S2).

Interruption is simulated to take place in the first year after the most recent survey for hypoendemic settings (i.e. year 1 for hypoendemic at baseline, year 4 for hypoendemic after 3 rounds of MDA and year 6 for hypoendemic after 5 years MDA), in the second year after most recent survey for mesoendemic settings, and in the third year for hyperendemic settings.

#### Impact of mitigation strategies

To evaluate in more detail the potential for mitigating the impact of COVID-19, we simulate both interruption and mitigation strategies in settings that would have been expected to reach the TF_1-9_ <5% threshold before 2030 with a strategy of annual district-level MDA targeting the whole community, but where an interruption is expected to cause a delay in achieving this control threshold (corresponding to an R_0_>1 and an R_T_<1; defined as MDA subcritical by Blumberg and colleagues^9^). Two settings with differing levels of transmission are considered: Setting 1 corresponding to a mean baseline (before MDA) TF_1-9_ of 40% (range 37.5-42.5), and Setting 2 corresponding to a mean baseline TF_1-9_ of 20% (range 17.5-22.5). A 12-month delay in MDA (yielding a 2-year gap in treatment) is simulated mid-way through a planned programme, assuming a 5-year MDA programme for Setting 1 (interruption in year 3) and a 3-year MDA programme for Setting 2 (interruption in year 2). Two mitigation strategies that could be implemented after resuming activities were then considered:

**Mitigation protocol 1 (M1)**, an additional round of community-wide MDA (all ages) delivered 6 months after the programme restarts;

**Mitigation protocol (M2)**, an additional round of MDA targeting only children aged 1-9 years delivered 6 months after the programme restarts.

### Mitigation and acceleration

A third setting, denoted Setting 3, is simulated to represent those districts which after more than 10 years of annual MDA, are still endemic (corresponding roughly to R_T_>1, defined as supercritical by Blumberg).^9^ This is done by simulating a higher level of transmission and also by filtering the stochastic simulations to include only those where TF_1-9_ is ≥10% after 10 annual rounds of MDA (further details Supplementary Data). MDA interruption is simulated to take place in year 11. As this setting is intended to be representative of those settings in which previous models indicate a new paradigm may be needed to achieve EPHP, two alternative strategies are simulated. These are:

**Mitigation and acceleration protocol 1 (MA1)**, which consists of two extra rounds of MDA targeting children aged 2-9 years only, delivered one week and 3 weeks after the normal annual community-wide MDA; and

**Mitigation and acceleration protocol 2 (MA2)**, which consists of 3 extra rounds of MDA targeting children aged 2-9 years at 3 month intervals following an annual community-wide MDA.

These are two strategies that have confirmed approval in Ethiopia (clinicaltrials.gov identifier NCT03523156 for MA1; NCT03335072 for MA2).^18^

A population of size 1000 was considered for all simulations which were run for 16 simulated years, except for Setting 3 where 23 years were simulated. Initial sets of stochastic simulations were filtered to give at least 1000 simulations for analysis based on criteria described (baseline prevalence or prevalence after a given number of MDA rounds). For Settings 1 and 2 (including mitigation simulations) to ensure simulations were representative of settings where TF_1-9_<5% would have been achievable by 2030, simulations which did not reach TF_1-9_ <5% were also removed and not considered for analysis. The impact of systematic non-adherence is also explored for Setting 3 by setting the adherence correlation parameter *ρ* to 0, 0.3 or 0.5. Confidence intervals are estimated as 95th centiles. All simulations were implemented in R version 3.6.3.

## Results

Figure 3 summarises the delay to programmes following a 1 year interruption across a range of endemic settings and levels of transmission, described by TF_1-9_ (x-axis) at the most recent survey, which may have been at baseline, after 3 years of MDA or after 5 years of MDA (y-axis; further results Supplementary Data Table S2). As expected, higher baseline prevalence levels (which correspond to higher transmission settings) are more impacted by a one year interruption to MDA in terms of delay to reaching the EPHP threshold. In settings where the baseline prevalence was more than 40%, or where TF_1-9_ has not been achieved after 3 or 5 years of annual MDA, the delay to achieving the EPHP threshold is estimated to be substantially longer than the interruption. Furthermore, where TF_1-9_ is greater than 15% after 3 years of MDA or greater than 10% after 5 years of MDA, the median time to achieving the threshold is longer than the length of the simulation (16 years).

**Figure 2.**
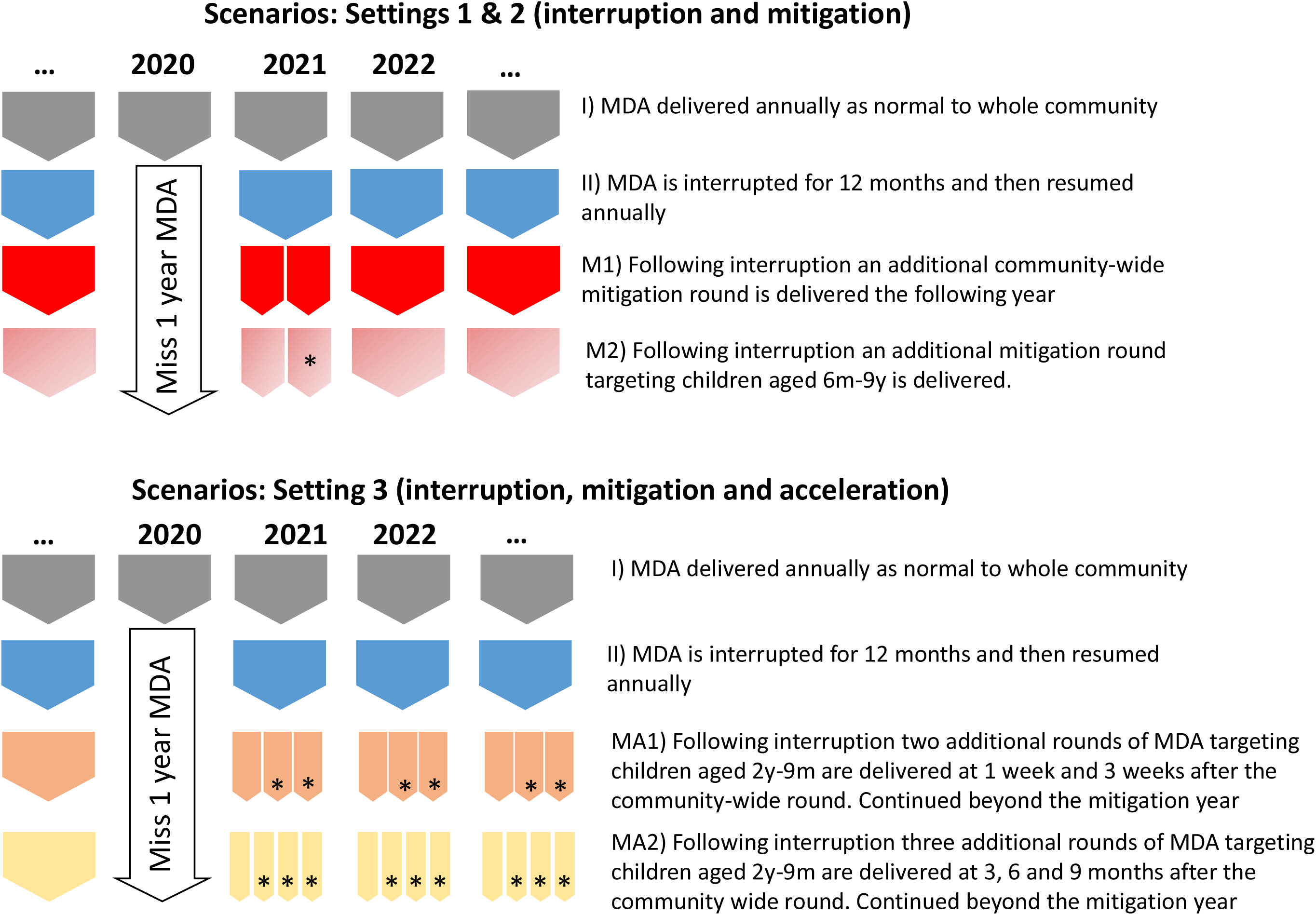
Simulated scenarios for Settings 1 and 2 (interruption and mitigation) and Setting 3 (interruption and mitigation/acceleration protocols). *denotes MDA round targeting children only.

**Figure 3.**
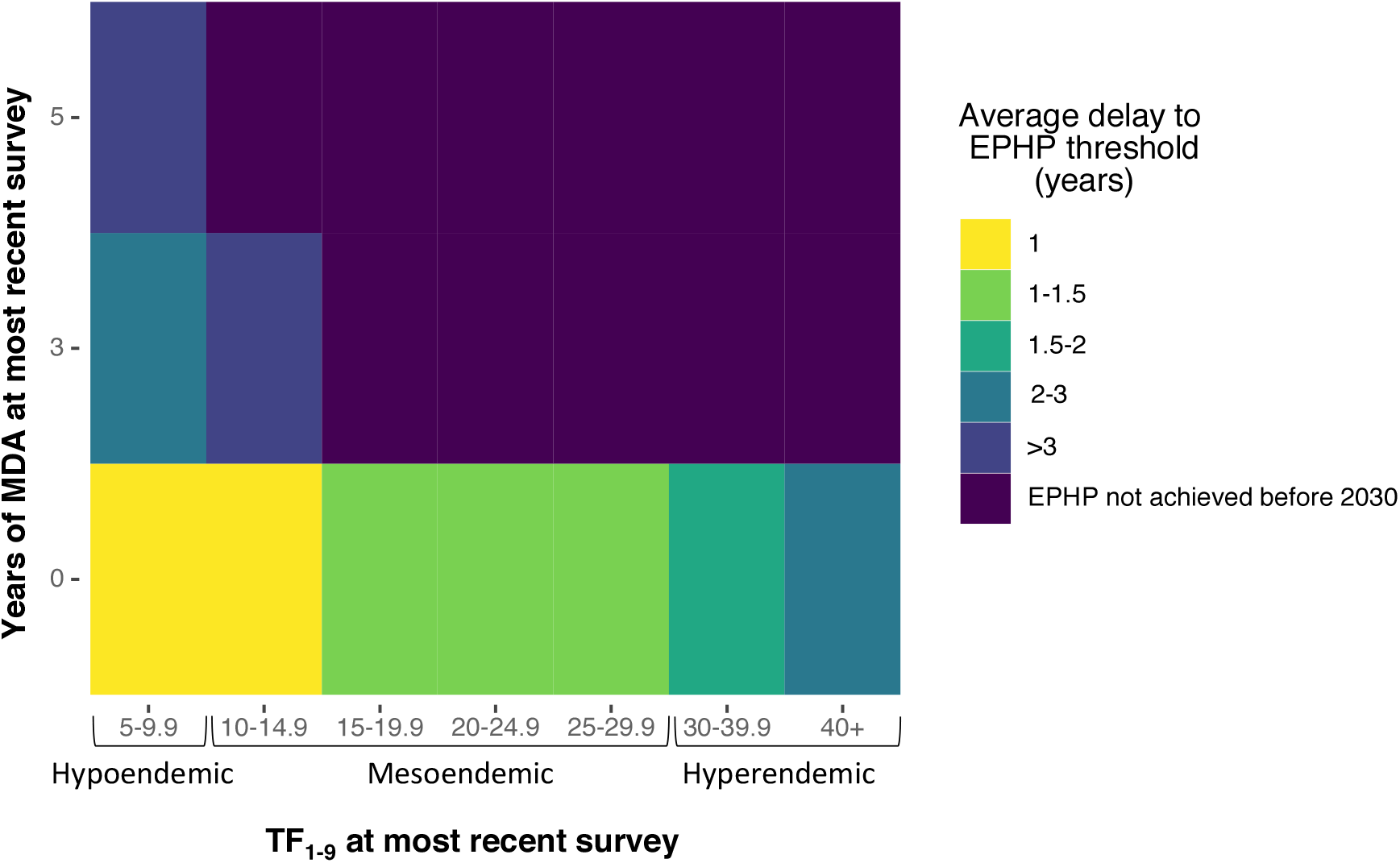
Median delay (years) to reaching EPHP threshold (TF_1-9_<5%) at varying levels of endemicity/stages of trachoma elimination programmes following a 1 year interruption to MDA. Years of MDA indicates the number of rounds of MDA that had been delivered when the last survey was carried out (0 representing baseline survey).

Figure 4 (panels B and D) shows that with an additional round of community-wide MDA in the year following interruption (mitigation scenarios M1), the prevalence of infection for both Setting 1 and Setting 2 is effectively reduced to where it would have been had the interruption not occurred. This has the effect of reducing the delay to achieving the EPHP threshold so that it is close to the duration of the interruption (12 months) for both settings (Figure 4 Panels A and B; Table 1), but due to the slower bounce-back rate the additional benefit of the mitigation round is not so pronounced in Setting 2.

**Table 1:** Summary of model output for Setting 1 and Setting 2. Confidence intervals are given as 95th centiles.

| **Setting 1** | **Mean years to achieve EPHP (Median; 95% CI)** | **% simulations reaching TF<sub>1-9</sub><5% after 6 rounds of MDA** | **Mean % TF in children after 6 rounds MDA (Median; 95% CI)** |
| --- | --- | --- | --- |
| **I) No interruption** | 4.4 (4.2; 2.4-11.5) | 88.1 | 1.7 (0; 0-12.9) |
| **II) 2020 interruption; No mitigation** | 7.1 (6.3; 2.4->16<sup>a</sup>) | 69.9 | 4.1 (1.5; 0, 19.9) |
| **M1) 2020 interruption; Extra MDA round: Community** | 5.3 (5.2; 2.4->16<sup>a</sup>) | 81.6 | 2.1 (0; 0-12.8) |
| **M2) 2020 interruption; Extra MDA round: Children** | 5.4 (5.3; 2.4->16<sup>a</sup>) | 77.9 | 2.8 (0.42; 0-15.2) |
| **Setting 2** | **Mean years to achieve EPHP (Median; 95% CI)** | **% simulations reaching TF<sub>1-9</sub><5% after 4 rounds of MDA** | **Mean % TF in children after 4 rounds MDA (Median; 95% CI)** |
| **I) No interruption** | 2.7 (2.6; 1.8-4.6) | 98.7 | 0.9(0.4; 0-4.2) |
| **II) 2020 interruption; No mitigation** | 4 (3.9; 1.8-5.7) | 97.1 | 0.9 (1.6; 0-5.2) |
| **M1) 2020 interruption; Extra MDA round: Community** | 3.8 (3.8; 1.8-4.6) | 99.2 | 0.5 (0; 0-5.8) |
| **M2) 2020 interruption; Extra MDA round: Children** | 3.9 (3.8; 1.8-4.9) | 99.2 | 0.5 (0; 0-5.8) |

<sup>a</sup>TF<sub>1-9</sub> is not reached within the timescale of the simulations (16 years).

**Figure 4.**
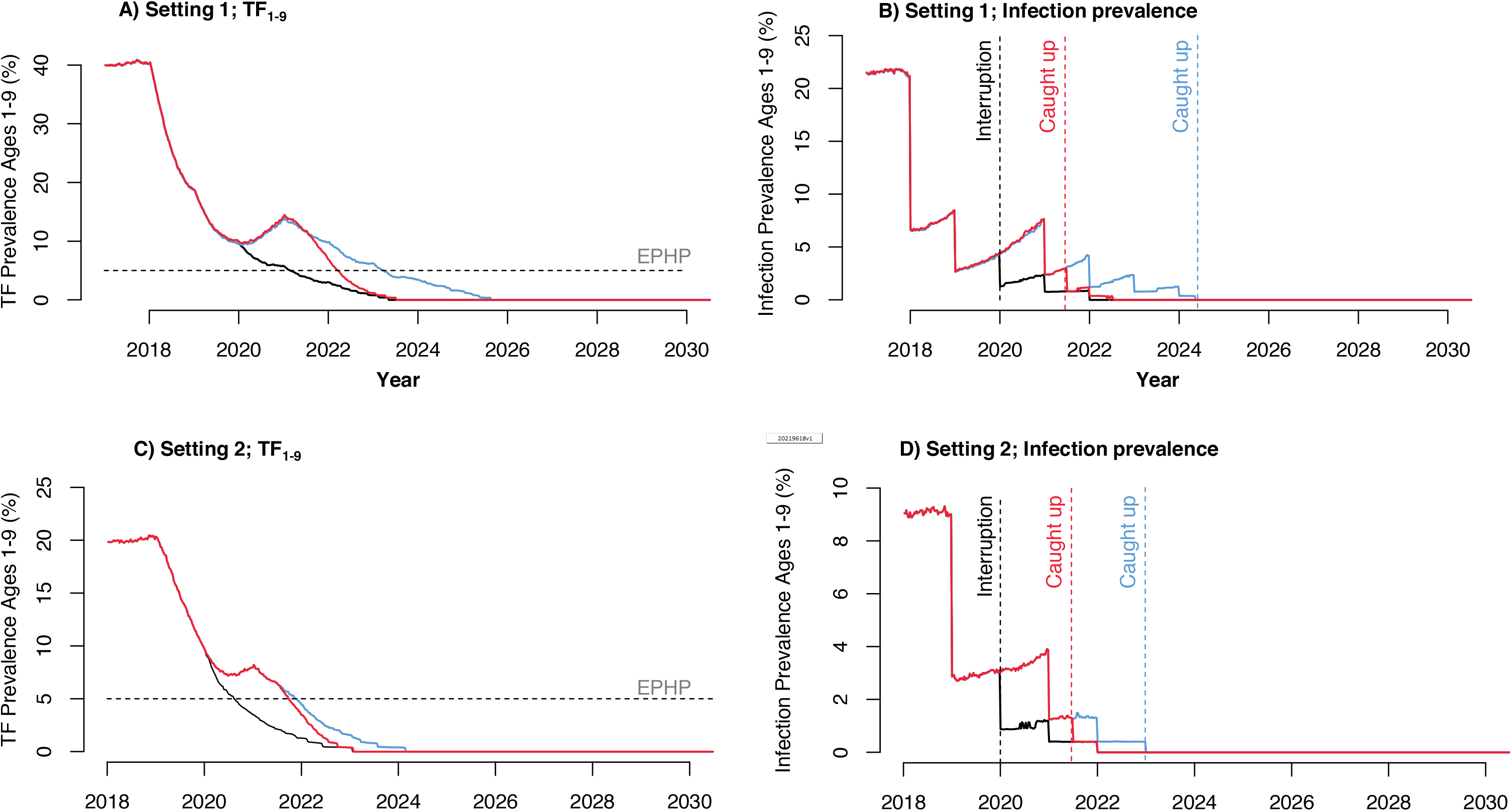
Median prevalence of TF in children aged 1-9 years (TF_1-9_) and ocular *Chlamydia trachomatis* infection in Setting 1 (Panels A and B) and Setting 2 (panels C and D). The black curve represents no disruption to MDA (scenario I in Figure 2), the blue curve represents a 1 year interruption with no mitigation (scenario II in Figure 2) and the red curve represents an additional mitigation round of community-wide MDA given in the year following interruption (scenario M1 Figure 2).

The impact of a mitigation round targeting children only (mitigation scenario M2; Table 2) in terms of delay and probability of reaching the threshold after a given number of MDA rounds is very similar to scenario M1 in which residents of all ages are offered antibiotics in the mitigation round.

**Table 2:**
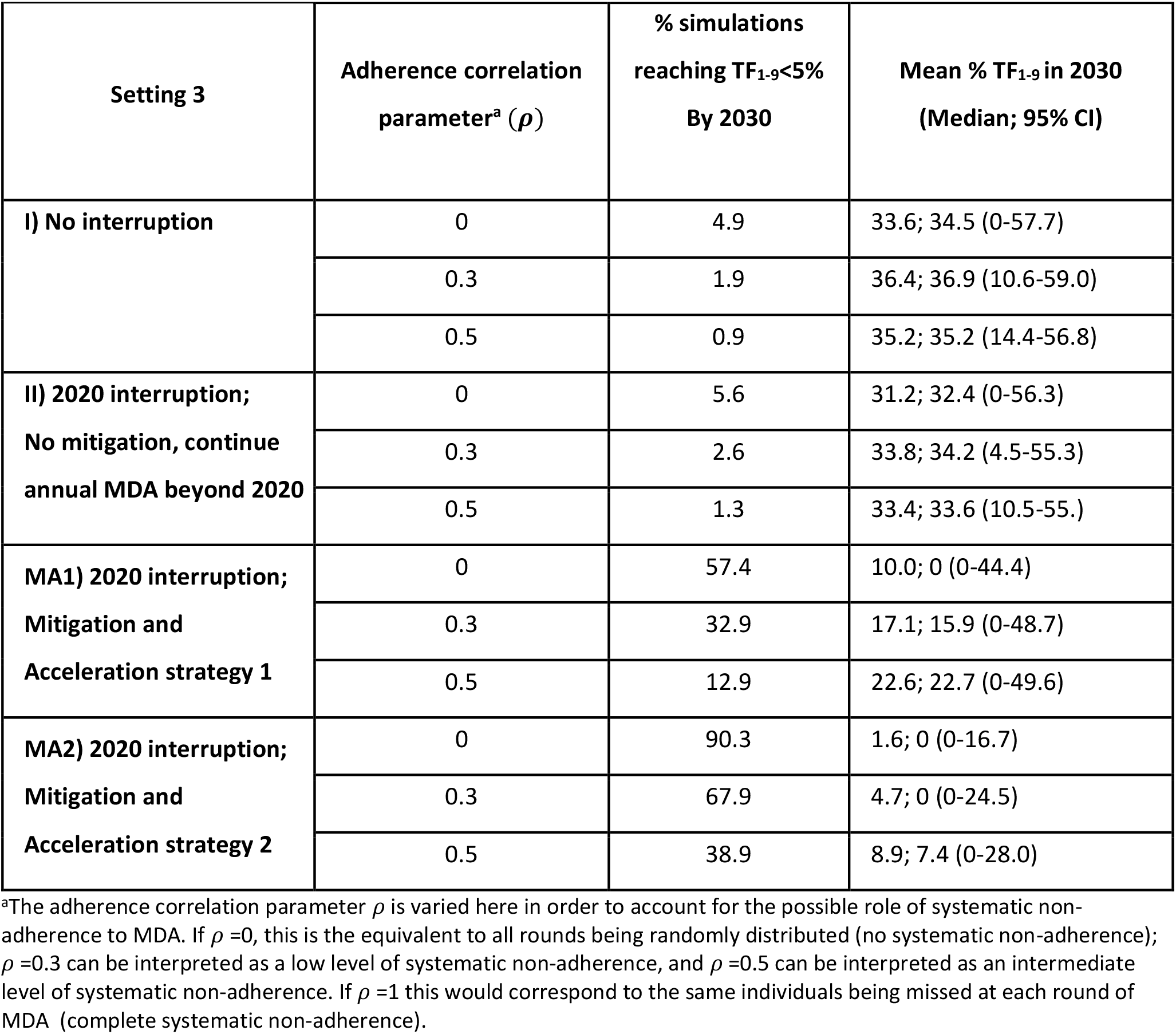
Summary of model output for Setting 3 (TF_1-9_>10% after 10 years of MDA). Confidence intervals are given as 95th centiles.

The simulations for Setting 3 (Figure 5 and Table 2) indicate that even without the interruption due to COVID-19 (scenario I), transmission levels in this setting are such that in the context of annual MDA, the probability of reaching the EPHP threshold for active trachoma is very low, even after 20 years of MDA and if no systematic non-adherence is assumed. In this Setting, only 4.9% of simulations reach TF_1-9_<5% by 2030 when treatment is assumed to be random (adherence-correlation parameter *ρ* =0), and only 0.9% if *ρ* =0.5. Both the mitigation and acceleration strategies simulated (MA1 and MA2) show clear improvement in the probability of reaching the EPHP goal by 2030. Where no systematic non-adherence is assumed (*ρ* =0), the median year in which TF_1-9_<5% would be reached is estimated to be 2027 for MA1 (representing 21 rounds of MDA post-2020) and 2024 for MA2 (after 14 rounds of MDA). These estimates are based on the assumption that these protocols are implemented in 2021 with the same minimum coverage level as before interruption, and continued until the TF_1-9_<5% threshold is achieved. For the mitigation and acceleration strategies MA1 and MA2, if systematic non-adherence is assumed to increase, the probability of reaching the threshold decreases, but remains markedly higher than that for continued annual MDA (Table 2).

**Figure 5.**
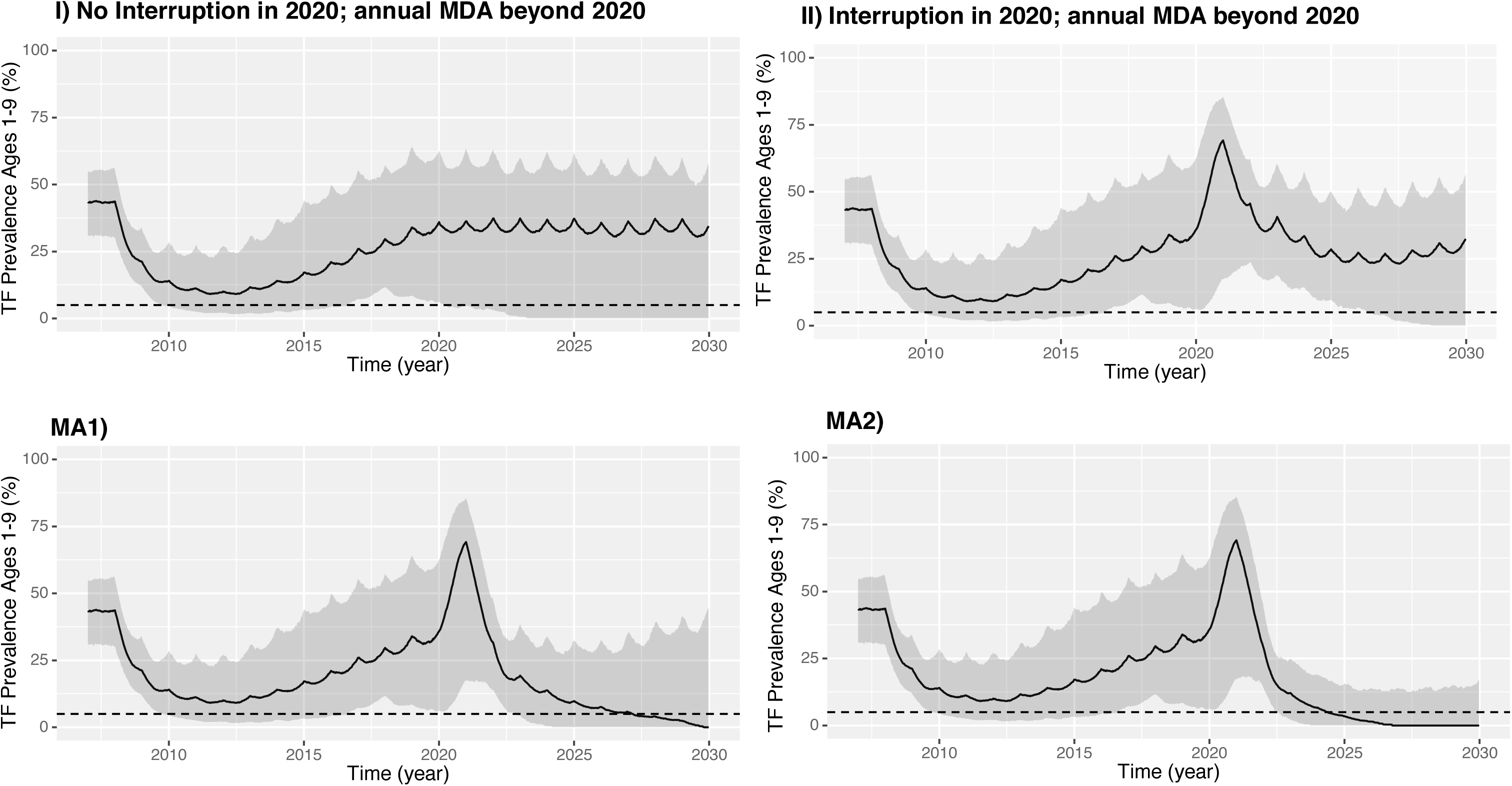
Median prevalence of TF in children aged 1-9 years (TF_1-9_) in Setting 3 (TF_1-9_>10% after 10 years of MDA). Adherence correlation parameter ρ r=0, corresponding to no systematic non-compliance. 95% confidence intervals are shown as shaded areas (estimated as 95^th^ centiles). MA1 and MA2 represent mitigation and acceleration strategies described in Figure 2.

## Discussion

Our results provide quantitative insights into the impact the current disruption to MDA activities may have on global efforts to eliminate trachoma, highlighting the need to prioritise mitigation strategies in those areas where the impact is predicted to be greatest. We also demonstrate the imperative need for alternative approaches in those areas where EPHP is unlikely to be achieved under existing protocols, notwithstanding the impact of COVID-19.

For those settings where EPHP would on average be achievable before 2030 (corresponding to R_T_<1 described by Blumberg and colleagues),^9^ but which were either hyperendemic at baseline (Setting 1) or where control has not been achieved after 3 years of MDA, our results indicate that missing a single round of treatment will lead to an increase in the number of MDA rounds needed to reach elimination targets. Following a 1-year interruption in these settings, it is estimated it will take on average more than 2 years to catch up if no mitigation protocols are implemented. By comparison, in Setting 2, where transmission is lower and baseline prevalence is mesoendemic (mean 20%), EPHP targets would still on average be reached after the same number of treatment rounds, and the length of the delay will be closer to the 1 year length of the simulated interruption. It is clear that as the level of transmission increases, the rate of resurgence during a period of programmatic interruption will also increase, which corresponds to the greater delay and number of treatment rounds necessary to make up lost ground once treatment is restarted. This is consistent with the expectation, supported by empirical evidence, that trachoma resurges more rapidly in higher prevalence settings.^7,8^ We also demonstrate that for both Setting 1 and Setting 2, a single additional round of MDA in the year following disruption will be sufficient to bring infection levels back to the level they would have been if the interruption had not occurred. The impact of this additional round is shown to be similar regardless of whether only children or the whole community are targeted. This is representative of the fact that in the model framework, in accordance with empirical evidence, children effectively act as a core group, due to both the higher bacterial loads and longer duration of infection that is assumed for early infections.^2,19–22^

It is noteworthy that for both Settings 1 and 2, even though mitigation MDA rounds bring infection levels back to where they would have been without interruption, this still leads to delay in achieving the EPHP threshold of approximately the same length as the interruption time period (just over one year). This is because the model represents the persistence of TF following resolution of infection. TF is a lagging indicator of infection at both the individual and population level, with correlation between TF and infection prevalence generally decreasing following MDA.^23^

With risk of donor fatigue a growing concern for many NTD control programmes,^24^ avoiding a prolonged delay to reaching EPHP targets is clearly a priority. Moreover, a delay to reaching trachoma thresholds will have a negative effect in terms of morbidity. Any surge in transmission during interruption, and every year of delay to achieving EPHP, corresponds to more *C. trachomatis* infections at both the individual and community level, the cumulative impact of which is more severe pathology and sequelae.^25^ In the mitigation scenarios explored here, additional catch-up rounds were simulated at 6 months after the programme restarted, and it is acknowledged that the timing of any additional rounds is likely to be primarily dictated by operational and financial constraints. Previous work suggests that regardless of timing, the anticipated mitigating effect of additional rounds of MDA delivered in the year following interruption would be equivalent.^26^ Therefore, to minimise impact both on morbidity and delay to achieving EPHP, additional rounds of MDA should be prioritised in high transmission settings, and delivered as soon as is practicable once programmes are able to restart at the same coverage levels as previously implemented.

Whilst a single additional round of MDA may be sufficient in some settings to bring infection levels back to where they would have been without interruption, for those settings where current schedules would not achieve EPHP even if interruption had not occurred, this is clearly far from adequate.

The mitigation and acceleration strategies simulated here represent two potential new paradigms of treatment for these settings, with both predicting clear improvement when compared to annual MDA. Both strategies have confirmed approval in Ethiopia, which may be similar to Setting 3. Whilst model predictions for MA2 indicate an increased probability of achieving EPHP compared to MA1, and trials have already demonstrated the efficacy of a similar protocol,^5^ logistical and financial constraints mean quarterly treatment (MA2) may be less feasible than three closely spaced treatment rounds (MA1). Furthermore, uncertainty regarding the assumptions of the model limits direct comparisons between the two protocols.

For both mitigation and acceleration strategies, if even a relatively low level of systematic non-compliance is assumed (*ρ* =0.3), the probability of achieving EPHP is decreased, although it would still be considerably greater than with annual MDA (at least a 10-fold increase in probability of achieving EPHP for either MA1 or MA2). However, the model assumes that individuals who are unlikely to adhere to treatment recommendations are randomly distributed throughout the population,^17^ whereas in reality, it is not only how many people are repeatedly missed by MDA, but who is missed, that determines the overall success of a programme. A study of trachoma endemic communities in Niger, for example, indicated that people who do not present for treatment are in fact less likely to be infected with ocular *C. trachomatis*.^27^ A study in Ethiopia indicated that levels of refusal were extremely low, at 0.6% of those offered azithromycin,^28^ with travelling during the campaign given as a major reason for non-treatment. Whether a protocol of repeated treatment rounds within a short space of time (MA1) or evenly spaced throughout the year (MA2) will maximise the probability of reaching all individuals targeted will likely be context-specific, but if systematically missed people are in fact less likely to be infected, the impact of non-adherence will be overestimated by the model. Another potentially important assumption of the model is that at the individual level, the infection-clearance outcome of receiving antibiotics is essentially binary and applied instantaneously. In reality however, even if an individual does not clear infection after treatment, their bacterial load is likely to be reduced. The implication of this is that the model predictions for MA1, in which three MDA rounds are delivered very close to each other, are likely to be somewhat pessimistic. Further empirical data on heterogeneity of bacterial load and efficacy of treatment for a given bacterial load would improve the biological realism of this aspect of the model.

Within the model framework used here, the higher transmission rates and baseline prevalence levels are simulated by increasing the transmission parameter. This parameter does not have a directly interpretable meaning in itself but can be considered a proxy for the range of factors that facilitate transmission of ocular *C. trachomatis* infection. These include overcrowding, lack of access to clean water, and comorbidities. Due to uncertainty regarding their impact, the model does not currently incorporate the potential reduction in transmission afforded by facial cleanliness and environmental improvement interventions, which also form part of the WHO strategy for trachoma control in addition to MDA.^15^ As such, the model predictions could be considered conservative. However, there are many potential indirect effects for COVID-19 on ocular *C. trachomatis* transmission in addition to the interruption to MDA which are as yet unknown. Given directives on physical distancing and the increased promotion of hygiene practices such as handwashing, it may be that transmission of *C. trachomatis* will decrease following COVID-19 in some settings. However, other possible consequences of COVID-19 such as migration, changes to health-seeking behaviour and economic hardship may exacerbate the increases in infection predicted by the model.

## Conclusion

The COVID-19 pandemic represents an unprecedented challenge to communities and healthcare systems worldwide. Whilst the current interruption to NTD control activities is clearly in line with the urgent need to minimise the spread of SARS-CoV-2, it is crucial that trachoma programmes are resumed at the same coverage levels as before interruption, as soon as is practicable. Further work is needed to confirm which districts should be prioritised for additional mitigation rounds of MDA once programmes are able to resume, and also to identify those districts where mitigation and acceleration are needed, with the current interruption potentially creating an opportunity to critically appraise and revise protocols as we look towards 2030.

## Data Availability

The datasets generated during and/or analysed during the current study are available from the corresponding author on request.

## Author’s contributions

The work in this manuscript was motivated by discussions prior to and during the 2020 technical meeting of the NTD Modelling Consortium. AB, JP and TDH conceptualised the study; AB designed the methodology and carried out the formal analysis; AB drafted the original manuscript; All authors edited and reviewed the manuscript.

## Disclaimer

The authors alone are responsible for the views expressed in this article and they do not necessarily represent the views, decisions or policies of the institutions with which they are affiliated.

## Acknowledgements

We would like to thank Robin Bailey and Amy Pinsent for helpful discussions whilst developing the model, and Simon Brooker, Paul Emerson and PJ Hooper for their valuable feedback on preliminary work.

## Supplementary Data

### Further details on transmission model

The model is based on a previously described framework^1^ which was validated and identified as the most parsimonious and best fit to cross-sectional PCR and TF data in a study comparing several possible frameworks for *C. trachomatis* transmission.^2^

The individual-based stochastic model described here incorporates some key aspects of ocular *C. trachomatis* infection biology, including acquired immunity leading to decreased duration of infection with repeated infection^3,4^ and allows simulation of some of the variability in response to MDA observed in empirical studies.^5,6^ Based on a modified SEIR framework (S=Susceptible; E=Exposed; I=Infectious; R=Recovered), individuals transition through four sequential states: Susceptible (S), infected but not yet diseased (I), infected and diseased (ID) or diseased but no longer infected (D). (See Figure 1 main text).

For each individual *i*, the duration of first ID and D periods (*ID*_*i,1*_; *D*_*i,1*_) are randomly assigned from Poisson distributions, with distribution means given as the baseline (longest) duration used by Pinsent and colleagues (see Table S1).^1^ The duration of these periods for subsequent infections are then assumed to decrease following a negative exponential to a minimum value, with decay rates and minimum durations also as given by Pinsent and colleagues.^1^ Similarly, it is assumed that an individual’s infectivity is proportional to their bacterial load, and that this also declines from the first infection following a negative exponential with each subsequent infection. For each individual’s (*i*) infection number (*j*), the calculated durations of *ID*_*i,j*_ and *D*_*i,j*_ are used as fixed transition periods, in contrast to exponential transitions utilised in the previous models.^1,7^ In order to ensure that the age-distribution of historical infections (and therefore infectivity, duration of infection and disease) are representative for a given level of transmission, a 40 year burn-in period was implemented for all simulations (burn-in period not considered for analyses).

To account for the possible role of systematic non-adherence to MDA, the “controlled correlation” method proposed by Dyson and colleagues is incorporated into the model.^8^ In this scheme, the first round of MDA is distributed randomly with probability given as the treatment coverage *c*. In each subsequent round *k*, individual *i* then receives treatment with probability *p*_*i,k*_ given as:

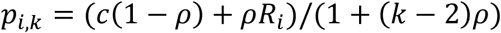

Where *R*_*i*_ is the total number of rounds attended by an individual previously, and *ρ* is the correlation parameter. If ρ =0, this is the equivalent to all rounds being randomly distributed (no systematic non-adherence) and if *ρ* =1 this corresponds to only those individuals who received the first round of treatment receiving future rounds (complete systematic non-adherence).

Test uncertainty and decision-making around reaching the TF_1-9_ threshold (which would normally determine whether MDA is halted) was not considered here.

**Table S1.** Model variables, parameters and sources.

| Notation | Description | Values /distribution | Units | Source |
| --- | --- | --- | --- | --- |
| $S_i$ | Susceptible individuals | - | | |
| $I_{i,j}$ | Infected and not yet diseased<br>(individual $i$ infection $j$) | 14 | Days | 9 |
| $ID_{i,1}$ | Infected and diseased period<br>(individual $i$, first infection) | $\sim \text{Poisson}(\omega_{max})$ | Days | |
| $ID_{i,j}$ | Infected and diseased period<br>(individual $i$, infection $j$) | $(ID_{i,1} - \omega_{min})e^{\phi(j-1)} + \omega_{min}$ | Days | 1,2,9 |
| $D_{i,1}$ | Diseased only period<br>(individual $i$, infection $j$) | $\sim \text{Poisson}(\tau_{max})$ | Days | |
| $D_{i,j}$ | Diseased only period<br>(individual $i$, infection $j$) | $(D_{i,1} - \tau_{min})e^{\theta(j-1)} + \tau_{min}$ | Days | 1,2,9 |
| $\omega$ | Duration of infected and diseased period | $\omega_{max} = 200, \omega_{min}=77$ | Days | 1,2,9 |
| $\tau$ | Duration of diseased only period | $\tau_{max} = 300, \tau_{min}=7$ | Days | 1,2,9 |
| $\phi$ | Decay rate, infected and diseased period | 0.45 | Proportion | 1,2,9 |
| $\theta$ | Decay rate, diseased only period | 0.3 | Proportion | 1,2,9 |
| $b$ | Infectivity of an individual proportional to their bacterial load | 0.114 | Proportion | 1,2 |
| $\beta$ | Transmission parameter | Varied to simulate a range of settings | | |
| $\lambda_a$ | Force of infection for age group $a$ | Calculated | Weeks | 1,2 |
| $\Gamma$ | Reduction in force of infection during disease only state | 0.5 | Proportion | 1,2,10 |
| $c$ | Treatment coverage (proportion of population receiving treatment at each round of MDA) | 0.8 | Proportion | 11 |
| $\epsilon$ | Treatment efficacy (probability of infection clearance given that treatment is received) | 0.85 | Proportion | 12 |
| $\rho$ | Compliance correlation parameter | 0, 0.3 and 0.5 considered | | 8 |

### Additional results: Estimating impact of 12 month interruption to MDA

Estimates are based on 1000 simulations for each category, filtered and sampled from an initial set of simulations to give a uniform distribution between category bounds.

Where the median/mean time to reaching the EPHP threshold without interruption to MDA is longer than the duration of the simulation (16 years), the delay cannot be estimated here and is given as NA.

**Table S2.** Summary of the impact of a 12 month interruption to MDA on reaching EPHP threshold of TF_1-9_<5% at varying levels of endemicity/stages of trachoma elimination programmes.

| Years of MDA, most recent survey | $TF_{1-9}$, most recent survey % | Median time to EPHP threshold; No interruption to MDA | Mean time to EPHP threshold; No interruption to MDA | Median time to EPHP threshold; 12 month interruption to MDA | Mean time to EPHP threshold; 12 month interruption to MDA | Median Delay to EPHP threshold | Mean Delay to EPHP threshold |
| --- | --- | --- | --- | --- | --- | --- | --- |
| 0 | 5-9.9 | 1.98 | 2.04 | 2.96 | 3 | 0.98 | 0.96 |
|  | 10-14.9 | 2.15 | 2.19 | 3.17 | 3.21 | 1.02 | 1.02 |
|  | 15-19.9 | 2.39 | 2.44 | 3.48 | 3.62 | 1.09 | 1.18 |
|  | 20-24.9 | 2.77 | 2.89 | 4.06 | 4.19 | 1.29 | 1.3 |
|  | 25-29.9 | 3.06 | 3.23 | 4.37 | 4.52 | 1.31 | 1.29 |
|  | 30-39.9 | 3.62 | 4.1 | 5.33 | 6.31 | 1.71 | 2.21 |
|  | 40+ | 4.6 | 5.31 | 7.15 | >16 | 2.55 | >9.69 |
| 3 | 5-9.9 | 4.6 | 4.77 | 7.06 | >16 | 2.46 | >10.23 |
|  | 10-14.9 | 6.75 | >16 | >16 | >16 | >8.25 | NA |
|  | 15-19.9 | >16 | >16 | >16 | >16 | NA | NA |
|  | 20-24.9 | >16 | >16 | >16 | >16 | NA | NA |
|  | 25-29.9 | >16 | >16 | >16 | >16 | NA | NA |
|  | 30-39.9 | >16 | >16 | >16 | >16 | NA | NA |
|  | 40+ | >16 | >16 | >16 | >16 | NA | NA |
| 5 | 5-9.9 | 7.31 | >16 | >16 | >16 | >7.69 | NA |
|  | 10-14.9 | >16 | >16 | >16 | >16 | NA | NA |
|  | 15-19.9 | >16 | >16 | >16 | >16 | NA | NA |
|  | 20-24.9 | >16 | >16 | >16 | >16 | NA | NA |
|  | 25-29.9 | >16 | >16 | >16 | >16 | NA | NA |
|  | 30-39.9 | >16 | >16 | >16 | >16 | NA | NA |
|  | 40+ | >16 | >16 | >16 | >16 | NA | NA |

### Additional results: Setting 3

An initial set of 4000 simulations was filtered to extract those where after 10 years of MDA, the prevalence of TF in children aged _1-9_ years (TF_1-9_) was more than 10%, giving a subset for analysis of at least 1000 simulations for each considered value of the adherence-correlation parameter *ρ* (higher values of *ρ* were not explored as they were considered unrealistic). Distributions of TF_1-9_ at baseline, and in 2030 are given in Figure S1, considering three possible strategies post-2020 (assuming a 2020 interruption to MDA). These are continuing annual MDA as previously, (scenario II in main text), and mitigation and acceleration strategies MA1 and MA2 as described in main text.

**Table S3.** Summary of Setting 3 at baseline. 95% Confidence intervals (CI) are given as 95^th^ centiles.

| | Adherence-correlation parameter<br>$\rho$ | Median prevalence<br>(95% CI) | Mean<br>prevalence (s.d) |
| --- | --- | --- | --- |
| Baseline | 0 | 43.5 (31.0,54.5) | 43.2 (6.1) |
|  | 0.3 | 43.3 (30.6,54.6) | 43.1 (6.32) |
|  | 0.5 | 42.0 (30.1, 54.7) | 42.9 (6.34) |

It is acknowledged that the baseline prevalence estimates for Setting 3 are likely to be lower than those observed in settings where control has not been achieved after more than 10 years of MDA. This is likely due to the fact that the initial model was fitted and parameterised using data from settings where rates of transmission is such that annual MDA has largely been successful.^2,3,9^ The implications of this are that some model assumptions (for example reduced infection/disease duration being a function of infection history and acquired immunity alone and not a combination of age and infection history) and parameter estimations are less representative of very high transmission settings. This means that in such settings, the non-linearity in the model of the relationship between the transmission parameter and baseline TF may mean baseline TF is underestimated. However the qualitative trends (equilibrium despite annual MDA, shift in this equilibrium with enhanced MDA) are still relevant. Further model fitting to relevant data is anticipated to improve the realism of the assumptions and parameter estimates for very high transmission settings.

**Figure S1.**
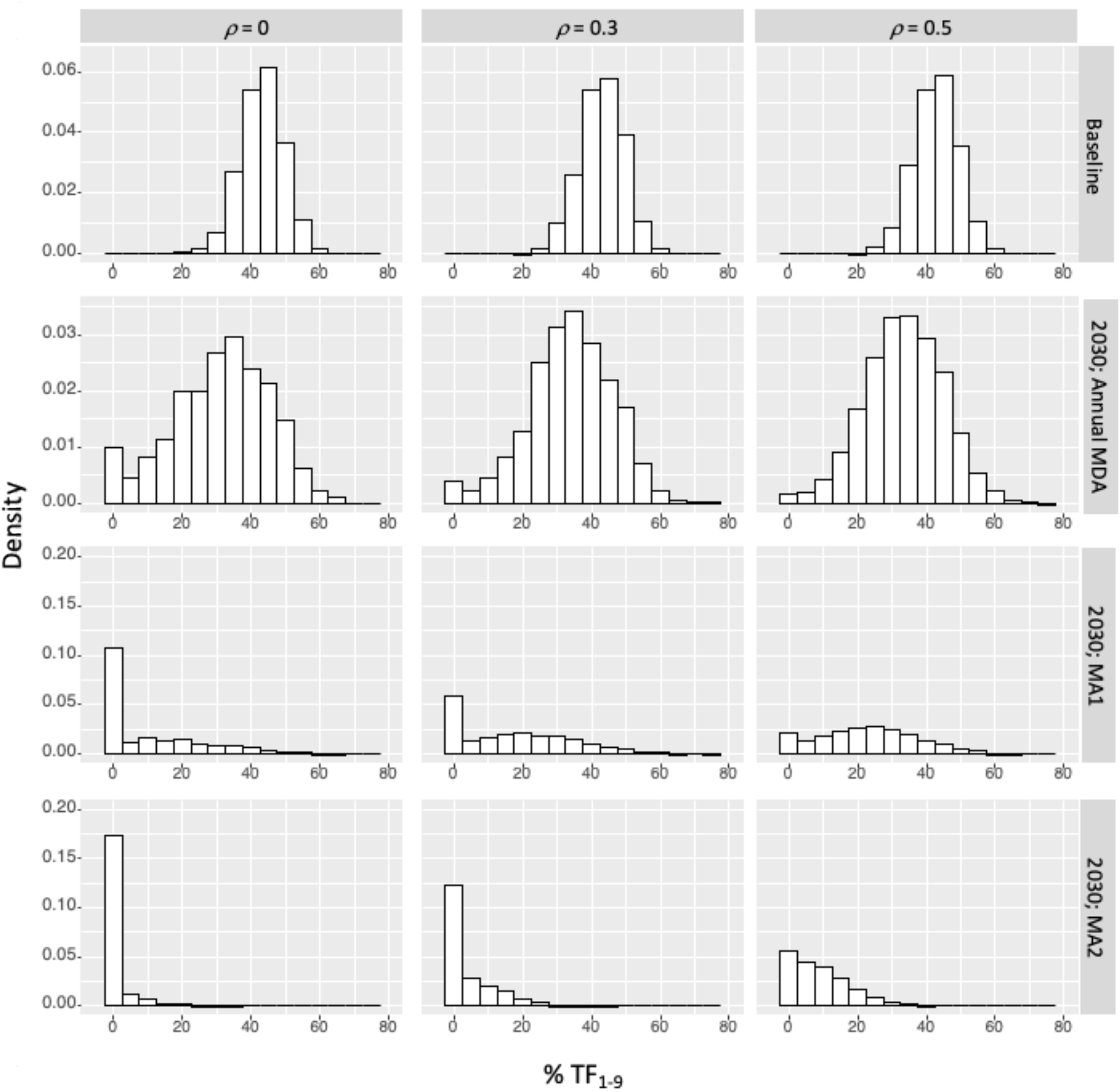
Distributions of TF prevalence in children aged 1-9 years (% TF_1-9_) at baseline (row 1) and in 2030 (rows 2-4). 2030 distributions follow simulation of either annual MDA beyond the 2020 interruption (row 2), or mitigation and acceleration strategies MA1 (row 3) or MA2 (row 4).

**Table S4.**
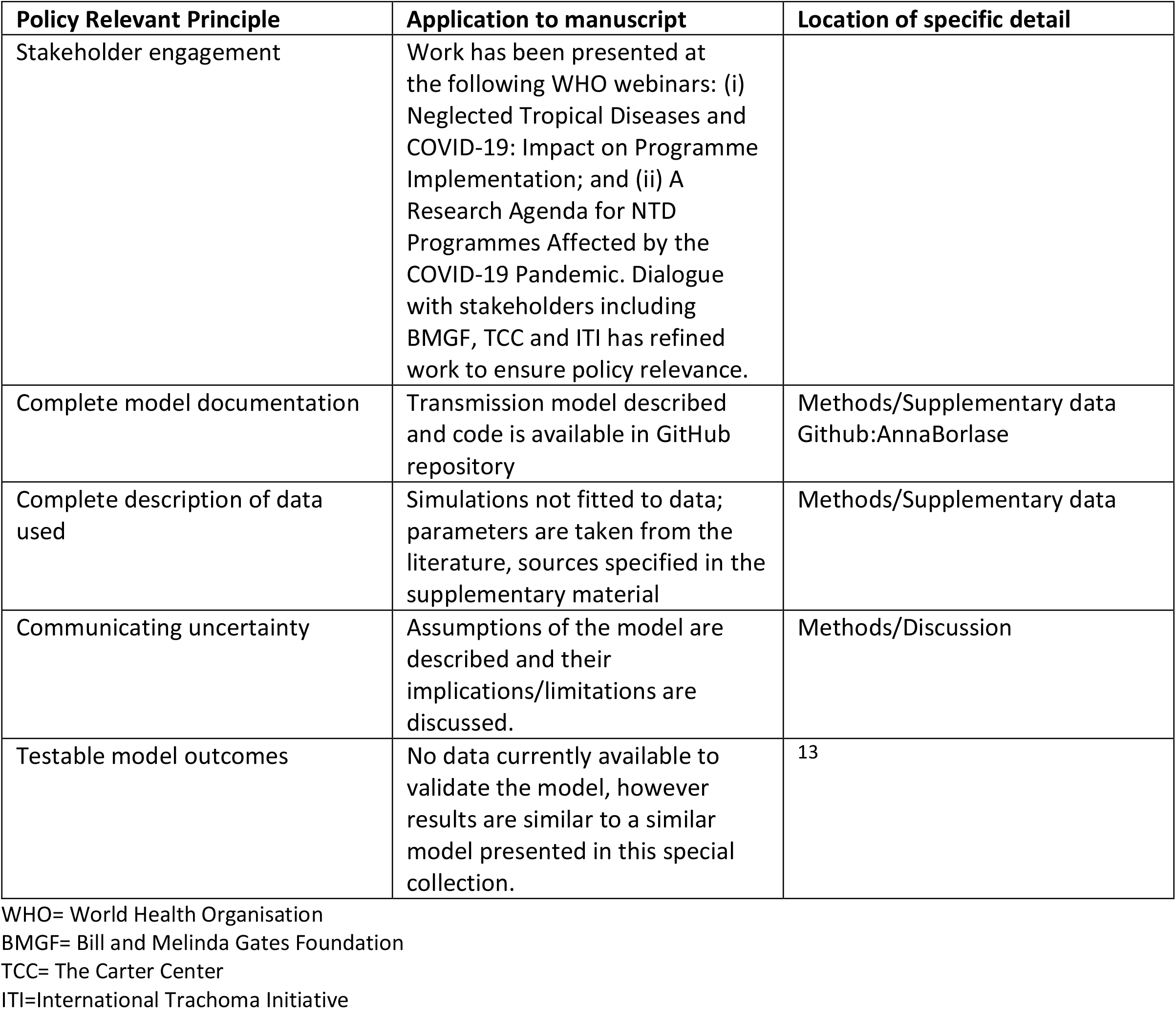
Policy-Relevant Items for Reporting Models in Epidemiology of Neglected Tropical Diseases (PRIME-NTD).^14^.

## References

1. World Health Organisation COVID-19 Interim Guidance. https://www.who.int/neglected_diseases/news/COVID19-WHO-interim-guidance-implementation-NTD-programmes/en/ (accessed 6th August 2020).

2. Bailey R, Duong T, Carpenter R, Whittle H, Mabey D. The duration of human ocular Chlamydia trachomatis infection is age dependent. Epidemiology and Infection 1999; 123(3):479–86. doi:10.1017/s0950268899003076

3. Schachter J, West SK, Mabey D, et al. Azithromycin in control of trachoma. Lancet (London, England). 1999; 354(9179):630–35. doi:10.1016/S0140-6736(98)12387-5

4. Chidambaram JD, Alemayehu W, Melese M, et al. Effect of a single mass antibiotic distribution on the prevalence of infectious trachoma. JAMA. 2006; 295(10):1142–46. doi:10.1001/jama.295.10.1142

5. House JI, Ayele B, Porco TC, et al. Assessment of herd protection against trachoma due to repeated mass antibiotic distributions: a cluster-randomised trial. Lancet (London, England). 2009; 373(9669):1111–18. doi:10.1016/S0140-6736(09)60323-8

6. Lietman TM, Oldenburg CE, Keenan JD. Trachoma: Time to Talk Eradication. Ophthalmology. 2020; 127(1):11–13. doi:10.1016/j.ophtha.2019.11.001

7. Lakew T, House J, Hong KC, et al. Reduction and return of infectious trachoma in severely affected communities in Ethiopia. PLoS Neglected Tropical Diseases. 2009; 3(2):e376. doi:10.1371/journal.pntd.0000376

8. Amza A, Kadri B, Nassirou B, et al. Effectiveness of expanding annual mass azithromycin distribution treatment coverage for trachoma in Niger: a cluster randomised trial. British Journal of Ophthalmology. 2018; 102(5):680–86. doi:10.1136/bjophthalmol-2017-310916

9. Blumberg S, Borlase A, Prada JM, Solomon AW, Emerson P, Hooper PJ, Deiner MS, Amoah B, Hollingsworth TD, Porco TC, Lietman TM. Implications of the COVID-19 pandemic on eliminating trachoma as a public health problem. Under Review. Published online 2020.

10. Lietman TM, Pinsent A, Liu F, Deiner M, Hollingsworth TD, Porco TC. Models of Trachoma Transmission and Their Policy Implications: From Control to Elimination. Clinical Infectious Diseases. 2018; 66(4): 275–80. doi:10.1093/cid/ciy004

11. Oldenburg CE, Amza A, Kadri B, et al. Comparison of Mass Azithromycin Coverage Targets of Children in Niger: A Cluster-Randomized Trachoma Trial. Am J Trop Med Hyg. 2018; 98(2):389–95. doi:10.4269/ajtmh.17-0501

12. Bailey RL, Arullendran P, Whittle HC, Mabey DC. Randomised controlled trial of single-dose azithromycin in treatment of trachoma. Lancet (London, England). 1993; 342(8869):453–56. doi:10.1016/0140-6736(93)91591-9

13. Pinsent A, Hollingsworth TD. Optimising sampling regimes and data collection to inform surveillance for trachoma control. PLoS Neglected Tropical Diseases. 2018; 12(10):e0006531. doi:10.1371/journal.pntd.0006531

14. Godwin W, Prada JM, Emerson P, et al. Trachoma Prevalence After Discontinuation of Mass Azithromycin Distribution. J Infect Dis. 2020; 221(Supplement_5):S519–S524. doi:10.1093/infdis/jiz691

15. World Health Organization. Report of the 3rd Global Scientific Meeting on Trachoma; 2010. Baltimore, USA.

16. Liu F, Porco TC, Mkocha HA, et al. The efficacy of oral azithromycin in clearing ocular chlamydia: mathematical modeling from a community-randomized trachoma trial. Epidemics. 2014; 6:10–17. doi:10.1016/j.epidem.2013.12.001

17. Dyson L, Stolk WA, Farrell SH, Hollingsworth TD. Measuring and modelling the effects of systematic non-adherence to mass drug administration. Epidemics. 2017; 18:56–66. doi:10.1016/j.epidem.2017.02.002

18. clinicaltrials.gov. Database of privately and publicly funded clinical studies around the world. (accessed August 5, 2020). clinicaltrials.gov

19. Nash SD, Chernet A, Moncada J, et al. Ocular Chlamydia trachomatis infection and infectious load among pre-school aged children within trachoma hyperendemic districts receiving the SAFE strategy, Amhara region, Ethiopia. PLoS Neglected Tropical Diseases. 2020;14(5):e0008226. doi:10.1371/journal.pntd.0008226

20. Solomon AW, Holland MJ, Burton MJ, et al. Strategies for control of trachoma: observational study with quantitative PCR. Lancet (London, England). 2003; 362(9379):198–204. doi:10.1016/S0140-6736(03)13909-8

21. West ES, Munoz B, Mkocha H, et al. Mass treatment and the effect on the load of Chlamydia trachomatis infection in a trachoma-hyperendemic community. Investigative Ophthalmology and Visual Science. 2005; 46(1):83–7. doi:10.1167/iovs.04-0327

22. Last A, Burr S, Alexander N, et al. Spatial clustering of high load ocular Chlamydia trachomatis infection in trachoma: a cross-sectional population-based study. Pathogens and Disease. 2017; 75(5). doi:10.1093/femspd/ftx050

23. Ramadhani AM, Derrick T, Macleod D, Holland MJ, Burton MJ. The Relationship between Active Trachoma and Ocular Chlamydia trachomatis Infection before and after Mass Antibiotic Treatment. PLoS Neglected Tropical Diseases. 2016; 10(10):e0005080. doi:10.1371/journal.pntd.0005080

24. World Health Organization. End the neglect to attain the sustainable development goals - A Road Map for Neglected Tropical Diseases 2021–2030; 2020.

25. Taylor HR, Burton MJ, Haddad D, West S, Wright H. Trachoma. Lancet (London, England). 2014; 384(9960):2142–52. doi:10.1016/S0140-6736(13)62182-0

26. Gao D, Lietman TM, Dong C-P, Porco TC. Mass drug administration: the importance of synchrony. Math Med Biol. 2017; 34(2):241–60. doi:10.1093/imammb/dqw005

27. Amza A, Kadri B, Nassirou B, et al. The easiest children to reach are most likely to be infected with ocular Chlamydia trachomatis in trachoma endemic areas of Niger. PLoS Neglected Tropical Diseases. 2013; 7(1):e1983. doi:10.1371/journal.pntd.0001983

28. Astale T, Sata E, Zerihun M, et al. Population-based coverage survey results following the mass drug administration of azithromycin for the treatment of trachoma in Amhara, Ethiopia. PLoS Negl Trop Dis. 2018;12(2):e0006270. doi:10.1371/journal.pntd.0006270

## Supplementary Data: References

1. Pinsent A, Hollingsworth TD. Optimising sampling regimes and data collection to inform surveillance for trachoma control. PLoS Neglected Tropical Diseases. 2018; 12(10):e0006531. doi:10.1371/journal.pntd.0006531

2. Pinsent A, Gambhir M. Improving our forecasts for trachoma elimination: What else do we need to know? PLoS Neglected Tropical Diseases. 2017; 11(2):e0005378. doi:10.1371/journal.pntd.0005378

3. Bailey R, Duong T, Carpenter R, Whittle H, Mabey D. The duration of human ocular Chlamydia trachomatis infection is age dependent. Epidemiology and Infection. 1999; 123(3):479–486. doi:10.1017/s0950268899003076

4. Bailey RL, Arullendran P, Whittle HC, Mabey DC. Randomised controlled trial of single-dose azithromycin in treatment of trachoma. Lancet (London, England). 1993; 342(8869):453–456. doi:10.1016/0140-6736(93)91591-9

5. Lakew T, House J, Hong KC, et al. Reduction and return of infectious trachoma in severely affected communities in Ethiopia. PLoS Neglected Tropical Diseases. 2009; 3(2):e376. doi:10.1371/journal.pntd.0000376

6. Oldenburg CE, Amza A, Kadri B, et al. Comparison of Mass Azithromycin Coverage Targets of Children in Niger: A Cluster-Randomized Trachoma Trial. Am J Trop Med Hyg. 2018; 98(2):389–395. doi:10.4269/ajtmh.17-0501

7. Godwin W, Prada JM, Emerson P, et al. Trachoma Prevalence After Discontinuation of Mass Azithromycin Distribution. J Infect Dis. 2020;221(Supplement_5):S519–S524. doi:10.1093/infdis/jiz691

8. Dyson L, Stolk WA, Farrell SH, Hollingsworth TD. Measuring and modelling the effects of systematic non-adherence to mass drug administration. Epidemics. 2017; 18:56–66. doi:10.1016/j.epidem.2017.02.002

9. Grassly NC, Ward ME, Ferris S, Mabey DC, Bailey RL. The natural history of trachoma infection and disease in a Gambian cohort with frequent follow-up. PLoS Neglected Tropical Diseases. 2008; 2(12):e341. doi:10.1371/journal.pntd.0000341

10. Shattock AJ, Gambhir M, Taylor HR, Cowling CS, Kaldor JM, Wilson DP. Control of trachoma in Australia: a model based evaluation of current interventions. PLoS Neglected Tropical Diseases. 2015;9(4):e0003474. doi:10.1371/journal.pntd.0003474

11. World Health Organization. Report of the 3rd Global Scientific Meeting on Trachoma. 2010. Baltimore, USA.

12. Liu F, Porco TC, Mkocha HA, et al. The efficacy of oral azithromycin in clearing ocular chlamydia: mathematical modeling from a community-randomized trachoma trial. Epidemics. 2014;6:10–17. doi:10.1016/j.epidem.2013.12.001

13. Blumberg S, Borlase A, Prada JM, Solomon AW, Emerson P, Hooper PJ, Deiner MS, Amoah B, Hollingsworth TD, Porco TC, Lietman TM. Implications of the COVID-19 pandemic on eliminating trachoma as a public health problem. Under Review. Published online 2020.

14. Behrend MR, Basáñez MG, Hamley JID, et al. Modelling for policy: The five principles of the Neglected Tropical Diseases Modelling Consortium. PLoS Negl Trop Dis. 2020; 14(4):e0008033. doi:10.1371/journal.pntd.0008033

